# From Awareness to ACTion Study: Improving Human Papillomavirus Knowledge, Screening, and Vaccine Uptake in Adolescent-Mother Pairs in the HOMINY study in Nigeria

**DOI:** 10.64898/2026.01.28.26344092

**Authors:** Oluwaseun Peter, Elohor Oborevwori, Esosa Osagie, Paul Akhigbe, Nosakhare L. Idemudia, Ozoemene Obuekwe, Fidelis E. Eki-Udoko, Nicholas Schlecht, Yana Bromberg, Nosayaba Osazuwa-Peters, Praise O. Okoh-Aihe, Modupe O. Coker, HOMINY Study Group

## Abstract

**Introduction:** Persistent infection with high-risk Human Papillomavirus (hr-HPV) in women is a leading cause of cervical cancer, and its co-infection among people living with HIV (PLHIV) increases the risk of HPV-associated cancer, including oropharyngeal and anogenital cancers. In sub-Saharan Africa, awareness of HPV is limited, screening and vaccine adoption are critically low, undermining efforts toward sexually transmitted infection (STI) elimination.

**Methods:** **F**rom **A**wareness to A**CT**ion (FACT) study assessed HPV knowledge, screening, and vaccine uptake in adolescent-mother pairs participating in the HOMINY (**H**PV, Human Immunodeficiency Virus, and **O**ral **M**icrobiota **I**nterplay in **N**igerian **Y**ouths) prospective cohort study. Participants were enrolled, including adolescents aged 9-18 years (N=636) and mothers aged 29-59 years (N=385). FACT was conducted at the University of Benin Teaching Hospital, Nigeria, with adolescent participants grouped as perinatally acquired HIV, HIV-exposed without acquisition and HIV-unexposed, and mothers by HIV serostatus. In line with the national immunisation programme protecting girls in Nigeria, sensitisation programmes were integrated into the research study to promote awareness and adoption of screening and vaccination practices. Knowledge and attitudes regarding HPV and its vaccination benefits were assessed through thematic discussions, field notes, interactive sessions, and questionnaires administered over the study period.

**Results:** At baseline, HPV awareness was low, with 34.5% of mothers and 1.4% of adolescents being aware of HPV. Post-sensitisation, awareness increased significantly to 64.4% and 19% in mothers and adolescents, respectively. Vaccination uptake rose from 0% to 3.4% in adolescents, and the proportion of mothers who underwent HPV-related screening (Visual Inspection with Acetic Acid and/or Papanicolaou test) increased from 38.7 % at baseline to 52.4 % after a year (p < 0.0001). Barriers to the adoption of preventive services included misconceptions, healthcare provider gaps, myths, misinformation, mistrust, skepticism, and limited access.

**Conclusions:** HPV awareness programmes significantly improved knowledge, vaccination uptake, and screening practices in this vulnerable population. As part of comprehensive STI elimination strategies, integrating HPV education and vaccination initiatives into HIV care and research will enhance prevention and address significant barriers. Lessons from a unique programmatic science framework provide critical insights for scaling vaccine delivery, and the design of future vaccine programmes.

## INTRODUCTION

Human Papillomavirus (HPV) is the most common viral, sexually transmitted infection worldwide and a leading cause of cervical cancer globally [1,2]. With over 95% of cervical cancer cases attributable to persistent HPV infection, it has been the predominant cause of Oropharyngeal Squamous Cell Carcinoma (OPSCC) in high-income countries, having surpassed tobacco and alcohol as the traditional cause [3,4]. The World Health Organisation (WHO) in 2022 estimated that the majority of sexually active individuals would acquire at least one type of HPV during their lifetime. While most HPV infections are short-lived and cleared spontaneously by the immune system, persistent infection with high-risk HPV types can lead to serious health conditions, including cervical cancer, OPSCC, other anogenital cancers, and genital warts [1].

In 2022, Nigeria recorded an estimated 13,676 new cervical cancer cases and 7,093 deaths, many of which are preventable through HPV vaccination and routine screening [2,5]. Although HPV poses a risk to the general population, people living with HIV (PLHIV), particularly women and adolescent girls, are disproportionately affected [5,6]. Their weakened immune systems heighten susceptibility to HPV infection and increase the likelihood of progression to malignancy [7]. With effective HPV vaccines available, awareness and uptake in Nigeria remain critically low [1,4]. Research has consistently revealed substantial knowledge gaps concerning HPV transmission modes, screening, risk factors, and vaccine safety within these populations [9,11,35].

In 2014, the World Health Organisation identified the HPV vaccine as a public health priority, recommending that it be included in the national immunisation programme. Nigeria, backed by GAVI in 2023, took a cue from this when it introduced and incorporated the single-dose Gardasil 4-Valent HPV vaccine, an Immunogen (Recombinant L1 proteins from HPV types): 6, 11, 16, 18 [4,10], into the national routine immunisation system for girls aged 9-14 years in October 2023 and May 2024 [11]. This marked a significant milestone in Nigeria’s efforts to eliminate cervical cancer, as the vaccine is 99% effective when administered [12]. The success of single-dose HPV vaccination strategies demonstrated in recent trials [13,14], combined with potential cross-protective benefits from other STI vaccines (e.g., meningococcal vaccines against gonorrhea), offers promise for streamlined, multipurpose STI prevention approaches [15,16]. Therefore, Nigeria’s introduction of single-dose HPV vaccination in 2023 presents an opportunity to accelerate progress toward the WHO’s cervical cancer elimination goals [12]. As sub-Saharan Africa bears a disproportionate burden of both HIV and HPV-related cancers, community-research partnerships like FACT provide valuable models for implementing evidence-based, contextually appropriate interventions that advance global STI elimination efforts.

Mothers and their children living with HIV, as well as the general population, exhibit a heightened vulnerability to persistent hr-HPV infection [6,7,20]. Therefore, it is crucial to assess their knowledge of HPV, screening practices, and vaccine uptake, while also identifying barriers to vaccination to guide targeted public health interventions [14,17]. In low- and middle-income countries (LMICs) in sub-Saharan Africa, where both HIV and HPV co-infection prevalence is not fully known and where the study is focused, integrating HPV education and immunisation strategies into HIV care and broader public awareness efforts could play a vital role in reducing the burden of cervical cancer and related diseases.

**F**rom **A**wareness to A**CT**ion (FACT) study, therefore, seeks to assess the level of HPV knowledge regarding screening, transmission modes, and vaccine awareness and uptake among adolescents and mothers enrolled in the parent prospective cohort study in Nigeria (HOMINY (**H**PV, Human Immunodeficiency Virus, and **O**ral **M**icrobiota **I**nterplay in **N**igerian **Y**ouths) study). HOMINY was aimed at evaluating the impact of the microbiome on the increased susceptibility to HPV [19,20,21]. The FACT study, in April 2024, systematically implemented a sensitisation programme through training and awareness. A month later, in May 2024, the second-phase five-day national vaccine drive in Benin City for girls aged 9-14 years was conducted, [22] followed by the full incorporation of the vaccine into national routine immunisation schedules for girls aged 9 years within health facilities nationwide. Additionally, FACT study also seeks to evaluate, longitudinally, the impact of the programme.

## METHODS

### Study participants

The FACT study implementation began at the 2^nd^ study visit of the parent study, HOMINY, which was a longitudinal HIV cohort study with a unique mother-adolescents population of PLHIV and groups without HIV acquisition, conducted at the University of Benin Teaching Hospital (UBTH), Benin City, Nigeria [21]. Participants were recruited from the paediatric special treatment HIV clinic for adolescents with perinatally acquired HIV (HI) on ART and HIV-exposed without acquisition (HEU) adolescents born to mothers living with HIV (HI), as well as from the general practice clinic for adolescents who are HIV-unexposed (HUU) and their mothers (HU). FACT participants were grouped into 385 mothers aged 29-59 years (262 HI and 123 HU) and 636 adolescents aged 9-18 years (220 HI, 203 HEU, and 213 HU), totaling 1,021. Participants were sensitised on HPV in April 2024, baseline data were collected and they were followed through three visits at 6-month intervals, from baseline to May 2025 1-year follow-up visit [21].

### Data collection procedure

This study employed a mixed-methods design, with quantitative and qualitative data guided by the Kirkpatrick Model [23], to evaluate the effectiveness of a research implementation and training program.

### Quantitative data

Data collection was conducted longitudinally across three of the four scheduled clinical visits during the HOMINY study period. A standardised and structured questionnaire was administered at each visit to comprehensively capture data relevant to HPV, with a particular emphasis on HPV screening, vaccine awareness, and uptake [20]. Detailed sociodemographic information was collected through validated, interviewer-administered questionnaires from both adolescents and their mothers or guardians. Variables assessed included age, sex, education level, income, and employment status. These factors were collected to evaluate their association with HPV exposure, health-seeking behaviour, and access to care [20,21]. Sexual health data were obtained using a confidential, age-appropriate questionnaire administered in a private setting by trained personnel. Items included sexual practices, condom use, and history of sexually transmitted infections (STIs). These data were critical for evaluating potential transmission pathways, particularly in relation to HPV. Medical history, including screening for HPV such as Visual Inspection with Acetic acid (VIA) and Papanicolaou smear (PAP), viral load for PLHIV, CD4/CD8, and vaccination status, was documented at each visit [21].

To maintain methodological consistency, all questionnaires were administered using a standardised protocol [21]. This mixed-methods approach combining clinical examination, medical record review, and structured interviews enabled a robust assessment of the complex interplay between social determinants, sexual behaviours, and infectious disease risk in the mother-adolescents population.

### Qualitative data

Sensitisation was provided to HOMINY study participants over a period of 3 weeks through thematic discussions and interactive sessions, with 60 mother-adolescent pairs per week as a subset of the entire population. Adolescents paired with their mothers were selected through criterion-based purposive sampling to ensure that all participants met the predefined age specifications for vaccination [24]. Nine trained study personnel with a combined experience of over 15 years coordinated each session, maintaining a ratio of 7 to 1. The model evaluated training outcomes across four hierarchical levels of the Kirkpatrick Model: reaction, learning, behaviour, and results, offering a comprehensive framework for understanding the programme’s impact [23]. The four sensitisation sessions addressed the first two levels of the model, reaction and learning [23]. In contrast, behaviour and results were evaluated using a blend of qualitative and quantitative data collected during the training and national vaccine rollout after the second assessment point [21,23].

In brief, the sensitisation programme was conducted in English and Pidgin English to make it more accessible and ensure participants understood and could contribute effectively. It comprised four sessions, each lasting approximately 15-20 minutes. We assessed the participants’ previous knowledge of HPV; trained them on HPV transmission modes, screening, and vaccination benefits; held interactive sessions to address questions from participants; and encouraged them to undergo screening within the facility where the research was ongoing, and to take advantage of the upcoming free vaccination program organised by the government. The facilitators took field notes of the responses (reactions and learning) in both English and Pidgin English; the latter was subsequently translated to English, and other team members reviewed these within 48 hours.

### Ethics

The ethical considerations for the study design were reviewed and approved by the institutional review boards at the Rutgers State University of New Jersey (Pro2022000949) and the University of Benin Teaching Hospital, Benin City (ADM/E22/A/VOL. VII/14813674), Nigeria. Informed consent and assent were obtained from all participants.

### Data analysis

Study data were collected and managed using REDCap electronic data capture tools hosted by University of Pennsylvania [25]. All analyses were conducted using R version 4.5.0, R Foundation for Statistical Computing, Vienna, Austria, and bar charts and tables were used to present the results [26].

## RESULTS

### Quantitative Results

A total of 636 adolescents (mean age 13.4 ± 2.5 years; range 9-18 years) were enrolled, comprising 220 HI, 203 HEU, and 213 HU participants who were age- and sex-matched. Alongside them, 385 mostly middle-aged mothers who attended at baseline, including 262 HI and 123 HU. As summarised in **Table 1**, there were no significant differences in adolescents’ sex distribution or in the ages of mother-adolescent pairs. Most participants had at least secondary education; 29.0% of mothers had tertiary education, while more than 50% of adolescents were still in secondary school. Among mothers living with HIV, 82.5% had a viral load <20 copies/ml, compared with 72.9% of ALHIV, with a high proportion of people living with HIV; 68.1% mothers and 34.6% adolescents, enrolled in the FACT study.

**Table 1:** Characteristics of Study Participants.

| Socio-Demographic characteristics - Mothers vs Adolescents |  |  |  |
| --- | --- | --- | --- |
| Adolescent (Characteristics) | HI<br>N = 220<sup>I</sup> | HEU<br>N = 203<sup>I</sup> | HUU<br>N = 213<sup>I</sup> |
| Age (years) |  |  |  |
| Mean ± SD | 13.7 ± 2.5 | 12.9 ± 2.3 | 13.6 ± 2.5 |
| Gender |  |  |  |
| Female | 113.0 (51.4%) | 108.0 (53.2%) | 113.0 (53.1%) |
| Male | 107.0 (48.6%) | 95.0 (46.8%) | 100.0 (46.9%) |
| Education |  |  |  |
| Primary | 84.0 (42.2%) | 75.0 (39.1%) | 60.0 (30.7%) |
| Secondary | 114.0 (57.3%) | 112.0 (58.3%) | 123.0 (65.1%) |
| Tertiary | 1.0 (0.5%) | 2.0 (1.0%) | 6.0 (3.2%) |
| Others | 0.0 (0.0%) | 3.0 (1.6%) | 0.0 (0.0%) |
| School Attended |  |  |  |
| Public | 110.0 (50.0%) | 118.0 (55.2%) | 106.0 (49.8%) |
| Private | 51.0 (23.2%) | 48.0 (23.6%) | 43.0 (20.2%) |
| NA | 59.0 (28.8%) | 37.0 (18.2%) | 64.0 (30.0%) |
| Viral load scale |  |  |  |
| <20 copies/ml | 159.0 (72.9%) | 0.0 (NA%) | 0.0 (NA%) |
| 20+ copies/ml | 59 (27.1%) | 0.0 (NA%) | 0.0 (NA%) |
| <sup>l</sup>n (%) |  |  |  |
| MOTHERS (Characteristics) | **HI**<br>N = 262<sup>l</sup> | **HU**<br>N = 123<sup>l</sup> |  |
| Age (years) |  |  |  |
| Mean ± SD | 44.2 ± 5.1 | 43.1 ± 5.6 | |
| Religion |  |  |  |
| Christian | 258.0 (98.5%) | 121.0 (98.4%) |  |
| Islam | 4.0 (1.5%) | 2.0 (1.6%) |  |
| Education |  |  |  |
| Primary | 72.0 (27.5%) | 14.0 (11.4%) |  |
| Secondary | 130.0 (49.6%) | 60.0 (48.8%) |  |
| Tertiary | 52.0 (19.8%) | 47.0 (38.2%) |  |
| Others | 8.0 (3.1%) | 2.0 (1.6%) |  |
| Income (Naira) |  |  |  |
| Mean ± SD | 47,576.3 ± 67,655.1 | 47,965.9 ± 45,413.9 | |
| Viral load scale |  |  |  |
| <20 copies/ml | 216.0 (82.4%) | 1.0 (100.0%) |  |
| 20+ copies/ml | 46.0 (17.6%) | 0.0 (0.0 %) |  |
| <sup>l</sup>n (%) |  |  |  |
| **Study group codes:** HI = Individuals (adolescents or mothers) living with HIV; HEU = adolescent HIV-exposed without acquisition; HUU = adolescent HIV-unexposed; HU = mothers HIV-unexposed. **\*Education:** Adolescent data used were collected at the 1-year study visit. |  |  |  |

In Table 2, awareness of HPV and its vaccine rose steadily across visits for both groups. Mothers’ HPV knowledge nearly doubled with an increase from 34.5% at 6 months to 64.4% at 18 months, and HPV vaccine awareness from 10.9% to 27.6% (p < 0.0001 and p = 0.023, respectively). Adolescents’ HPV knowledge improved from 1.4% to 19% and vaccine awareness from 0.3% to 12.8%; only the HPV knowledge trend was significant, p < 0.0001, vaccine p = 0.392. Gains were consistently greater among mothers than among adolescents.

**Table 2.** HPV knowledge and vaccine awareness.

| **Respondent** | **Visit** | **N** | **HPV knowledge (%)** | **HPV vaccine awareness (%)** | **P-value trend HPV knowledge** | **P-value trend vaccine awareness** |
| --- | --- | --- | --- | --- | --- | --- |
| Mothers | Baseline | 385 | 133 (34.5) | 42 (10.9) | <0.0001 | 0.023 |
| Mothers | 6-month visit | 359 | 176 (49.0) | 66 (18.4) | <0.0001 | 0.023 |
| Mothers | 1-year visit | 340 | 219 (64.4) | 94 (27.6) | <0.0001 | 0.023 |
| Youths | Baseline | 636 | 9 (1.4) | 2 (0.3) | <0.0001 | 0.392 |
| Youths | 6-month visit | 597 | 52 (8.7) | 43 (7.2) | <0.0001 | 0.392 |
| Youths | 1-year visit | 580 | 110 (19.0) | 74 (12.8) | <0.0001 | 0.392 |

As shown in Figure 1, specific knowledge about the transmission modes of HPV was significantly low, with only 0.3% and 0.5% of mothers reporting that HPV can be transmitted vertically and through kissing, respectively. No adolescents responded positively to being aware of either mode of transmission. However, this was not the case for sexual transmission, where a steady increase in awareness was observed across visits. The most significant improvement was seen in knowledge about HPV being causally linked cancer, with awareness among mothers rising from 33.2% to 63.8%, and among adolescents from 1.1% to 18.1%.

**Figure 1.**
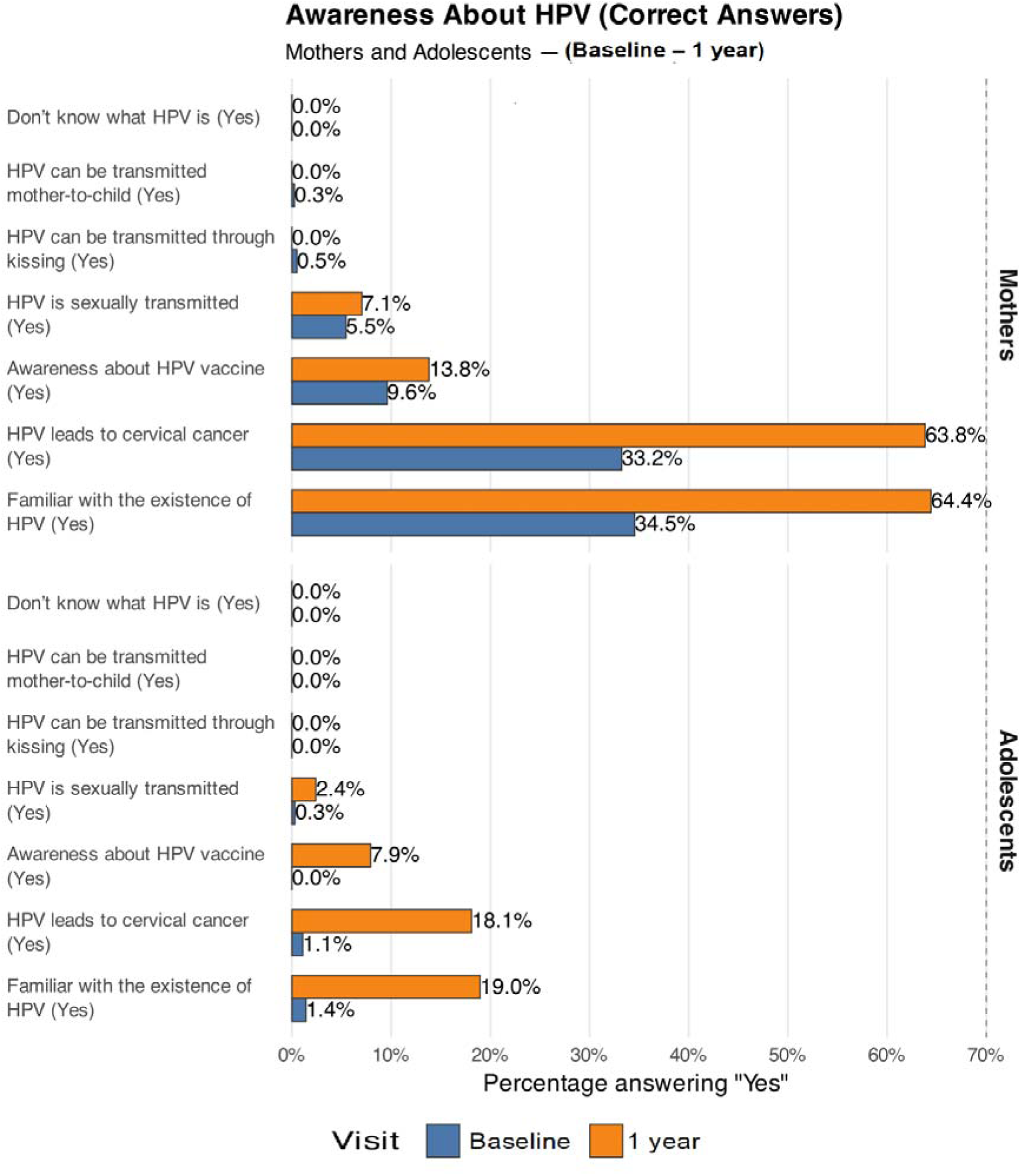
Awareness level about HPV.

A positive upward trend in HPV knowledge across visits for both groups was observed in Figure 2, with mothers consistently higher, highlighting the mothers’ stronger retention and comprehension in this older and more experienced group (i.e, mothers vs adolescents).

**Figure 2.**
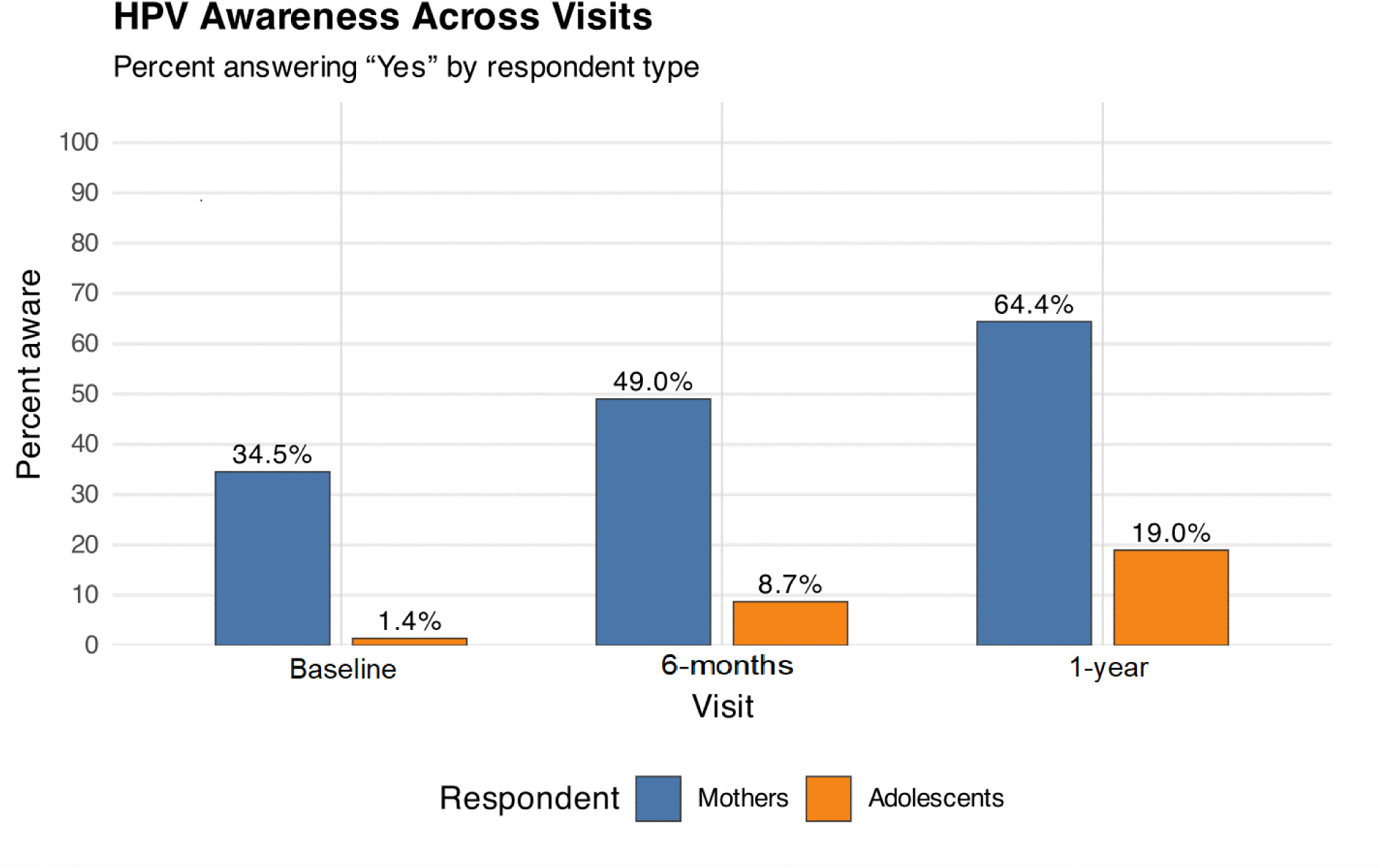
HPV awareness in mother-adolescent dyads across visits.

As shown in Table 3, VIA screening rose steadily from 35.6 % at 2^nd^ visit to 49.7 % at final visit (< 0.0001), indicating a highly significant upward trend. The PAP test results remained low and stable (≈ 10–11%) with a non-significant p-value of 0.649. There was an increase in any screening (VIA or PAP) from 38.7 % to 52.4 %, (p < 0.0001). Overall, this showed a marked and statistically significant increase in cervical screening, driven almost entirely by the growth in VIA participation, which was the primary test. In contrast, PAP smear screening rates remained unchanged.

**Table 3.** Screening across visits (Mothers only) - with trend p-values.

| Visit | N | VIA (%) | PAP (%) | Any screening (%) | VIA p-values | PAP p-values | Any screening p-values |
| --- | --- | --- | --- | --- | --- | --- | --- |
| Baseline | 385 | 137 (35.6) | 39 (10.1) | 149 (38.7) | $< 0.0001$ | 0.649 | $< 0.0001$ |
| 6-month visit | 359 | 150 (41.8) | 38 (10.6) | 162 (45.1) | $< 0.0001$ | 0.649 | $< 0.0001$ |
| 1-year visit | 340 | 169 (49.7) | 38 (11.2) | 178 (52.4) | $< 0.0001$ | 0.649 | $< 0.0001$ |

Table 4 presents the vaccine uptake in adolescents. At baseline, prior to sensitisation and national vaccine rollout, uptake stood at 0%; however, 6- and 12-months post-sensitisation, there was a consistent rise in uptake of the HPV vaccine to 2.3% and 3.4% respectively (p < 0.0001).

**Table 4.** Vaccine uptake across visits (Adolescents only) - with trend p-values.

| Visit | N | HPV vaccine dose 1 (%) | Dose p values |
| --- | --- | --- | --- |
| Baseline | 636 | 0 (0) | < 0.0001 |
| 6-month visit | 597 | 14 (2.3) | < 0.0001 |
| 1-year visit | 580 | 20 (3.4) | < 0.0001 |

VIA and/or PAP cervical screening tests done by mothers across the visits were illustrated in Figure 3. There was a consistent upward trend in screening for cervical cancer in the mothers, post-sensitisation

**Figure 3.**
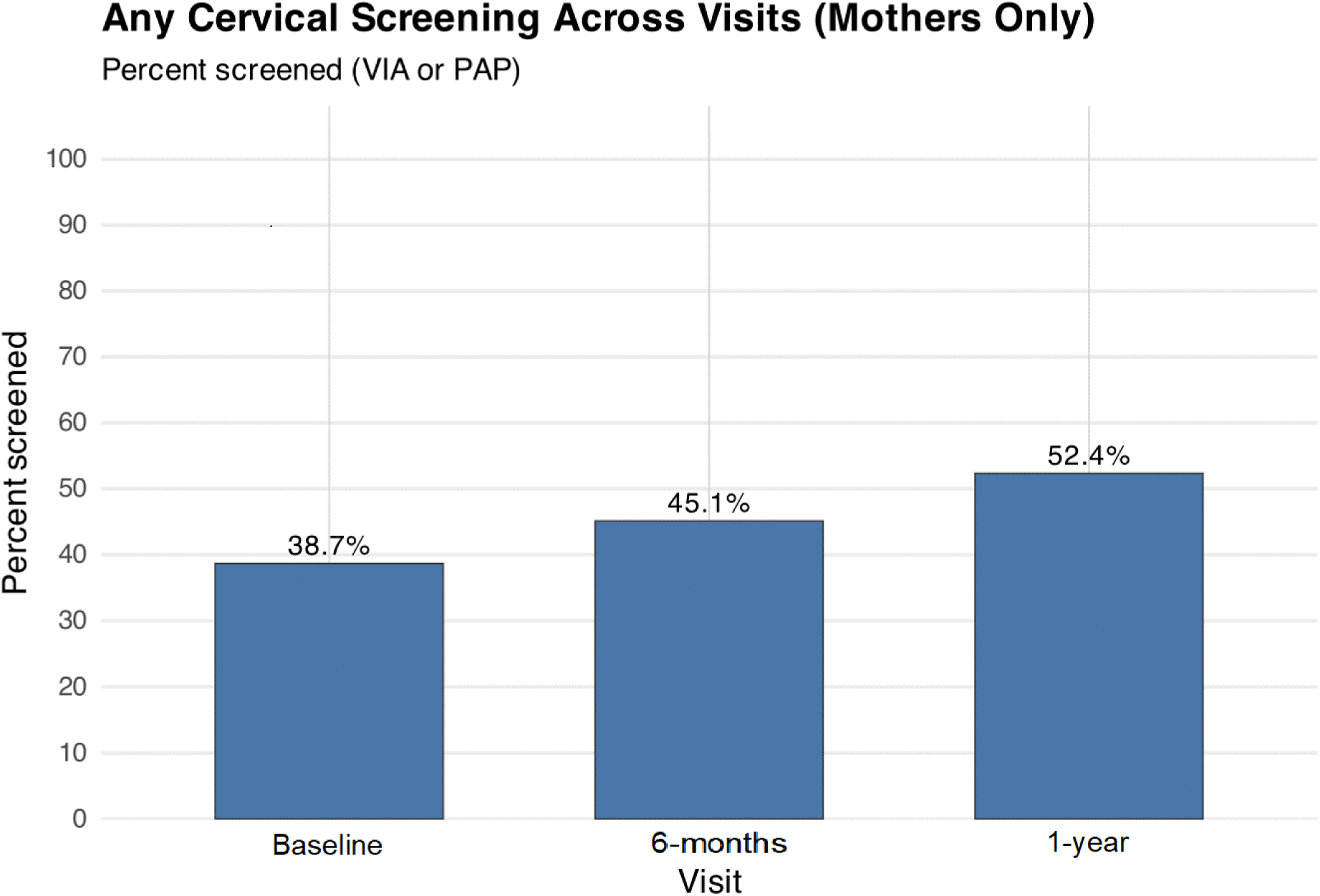
Any cervical screening (VIA or PAP) across visits in mothers.

The vaccine uptake trend in adolescents, depicted in Figure 4, shows a steady rise in vaccine uptake across visits, indicating the impact of the national free vaccination scheme and training.

**Figure 4.**
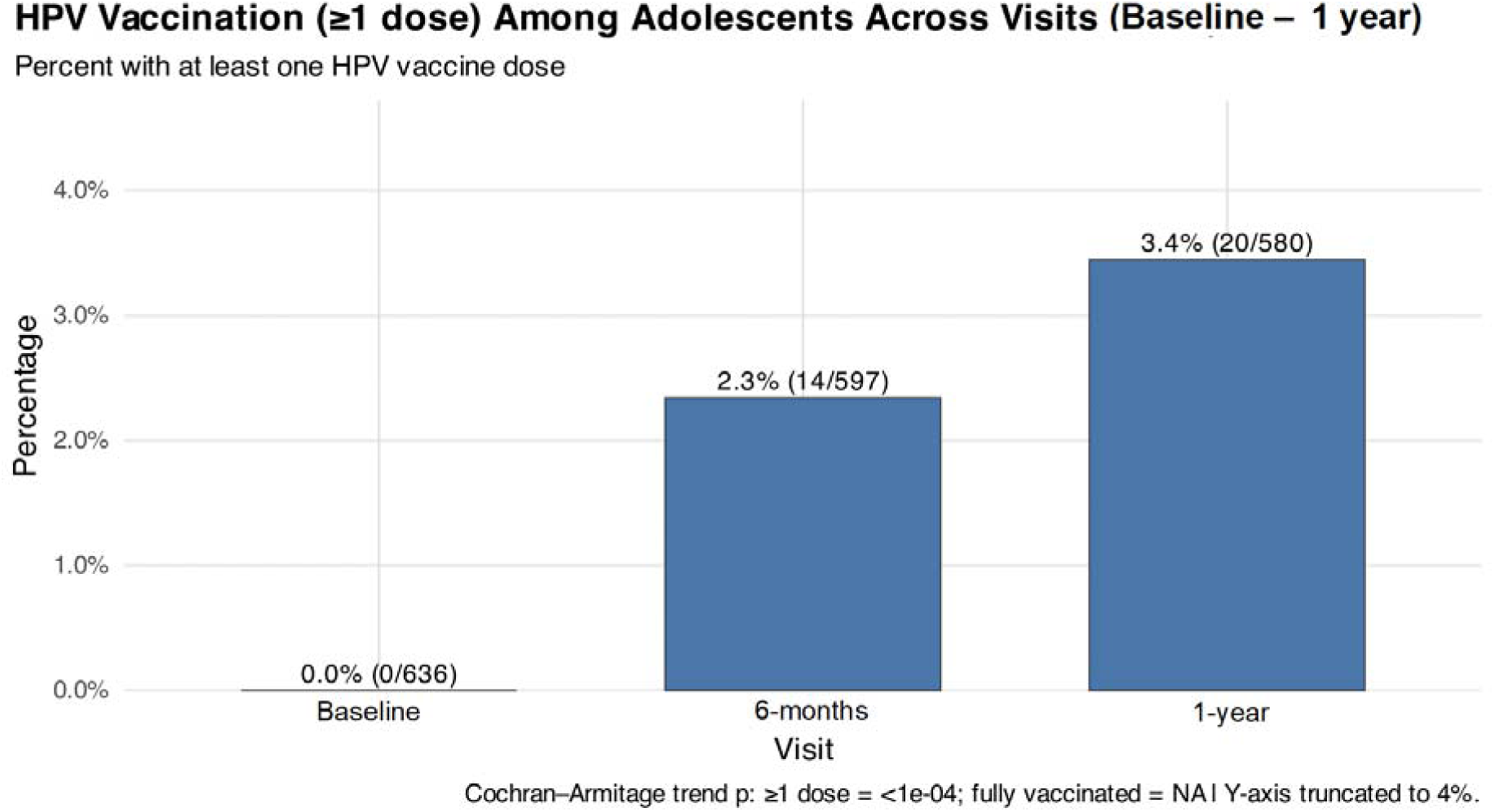
HPV vaccine uptake in adolescents at the baseline and in 6-months and 1-year post-sensitisation, respectively.

Table 5 shows that the main barrier to screening among mothers was a lack of perceived need when asymptomatic, reported by 60.5 % for VIA and 71.7 % for PAP testing. Another notable barrier was the absence of a health care provider’s recommendation, while smaller proportions cited misconceptions or limited knowledge.

**Table 5.** Barriers to Mothers’ screening at baseline (counts & percentage)

| Barrier | n | VIA % | n | PAP % |
| --- | --- | --- | --- | --- |
| Haven't had any problems | 150 | 60.5 | 248 | 71.7 |
| Doctor didn't order it/didn't say I needed it | 59 | 23.8 | 67 | 19.4 |
| Put it off/didn't get around to it | 15 | 6.0 | 4 | 1.2 |
| Didn't need it/didn't know I needed this type of test | 4 | 1.6 | 5 | 1.4 |
| Had hysterectomy | 2 | 0.8 | 2 | 0.6 |
| Don't Know | 18 | 7.3 | 20 | 5.8 |

Association between HPV knowledge and sexual practices among mothers from the baseline to the 4^th^ visit. Prevalence ratios with 95% confidence intervals (CIs) from Poisson regression models are shown on a logarithmic scale. Both unadjusted and adjusted estimates (for visit, age, and education) clustered around unity, with CIs crossing the null line, indicating no significant association between HPV knowledge and reported sexual practices.

Association between HPV knowledge and sexual practices among mothers, stratified by HIV serostatus across the study period. Both unadjusted and adjusted prevalence ratios clustered around 1, with confidence intervals crossing the null, indicating no significant association.

### Qualitative Results

The 180 mother-adolescent pairs (≈18%; 60 per week) selected for the qualitative study from the total study population of 1,021 share the same sociodemographic characteristics as presented in Table 1.

### Misconceptions

One of the commonly cited explanations for the failure to be screened in mothers was that awareness is relatively high, but knowledge of its importance is low, even though VIA is available free of charge in the facility. Some are unaware that screening is free, while others are not aware that the vaccine is more effective in adolescents aged 9-14 years.

> *“… I know that the test is free at UBTH but I don’t know why I should do it since I don’t have any health challenge.”*

> *“Can I take the vaccine or is it only for my daughter’s benefit?”*

> *“…I can’t have cancer, never! We don’t inherit cancer in our family…”*

Some do not even have the knowledge, nor are they aware of any such thing as cervical cancer, HPV being the cause, or that it can be prevented by vaccination.

> *“… I don’t know anything called HPV, I have only heard about HIV. What is this one [HPV] again?”*

### Vaccine mistrust

Some of the FACT study participants were aware of the vaccination; however, they believed that their children did not need it, and others were afraid of potential side effects, especially because it mainly targeted females. Parents/guardians did not think that boys were also at risk of acquiring HPV and were eligible for the vaccination. However, the national programme did not include boys in the vaccination scheme. The various conspiracy theories surrounding COVID-19 vaccination also played a significant role in shaping attitudes towards the vaccine uptake.

> *“This vaccine seems to be targeted at Nigeria to reduce our population. I cannot allow my girls to take it…”*

> *“…why are they giving vaccine to our girl child? Don’t they want them to get pregnant in the future… I don’t want trouble for my girl child after she gets married and is not able to conceive… over my dead body.”*

> *“I hope this one [vaccine] will not be worse than COVID vaccine, I could not move my hand for some time after I took the vaccine.”*

### Health care providers’ gaps

Another common observation was a lack of knowledge that HPV is responsible for cervical, oropharyngeal, and/or anogenital cancers and transmission modes of HPV, despite some mothers having undergone screening before the implementation of the FACT study. They believed that they were not at risk of cervical cancer and did not even know how it could be contracted.

> *“We were told during our clinic visit that it was now part of the test we need to do because we’re living with HIV, but we did not know why, and what the cause was.”*

> *“…the doctor just pointed to one of the rooms where the nurses were, he said I should do one test [VIA] and they put one liquid inside my private part, to check if I have HPV… I don’t know what causes it but after the test, I was told I don’t have HPV and that the test was negative.”*

> *“How can we protect or prevent ourselves from this infection? Maybe we can use protection [condom]…?”*

Most of the mothers who did the screening were PLHIV, with only a few knowing the test they were told to do in their clinic.

> *“I was told it is caused by a virus, and I even did what is called PAP smear to be sure I did not have it [cervical cancer] …”*

### Limited Access

Some participants were interested in undergoing the test and wanted their children to take the vaccine once rollout begins, with concerns about location and cost implications of the screening and vaccination.

> *“… please when will they start giving the vaccine so that my child can be vaccinated… how much will it cost or will it be free?”*

> *“Where in UBTH will I be able to do the screening, or can we do it in any hospital or primary health care centre close to us and how much is it?”*

> *“This test [VIA], how many days will it take to be ready, can my child be tested for it [VIA]?”*

## Discussion

This prospective FACT cohort highlights the persistent gap in awareness of HPV, its vaccine uptake, and screening in Nigeria, while demonstrating that targeted sensitisation can drive measurable improvements. Table 1 shows the socio-demographic characteristics of mothers and adolescents enrolled in the FACT study. At baseline, as shown in Table 2, only one-third of mothers and less than 2% of adolescents had heard of HPV, and vaccine awareness was even lower (10.9% of mothers; 0.3% of adolescents). Similarly, in 2020, HPV knowledge and vaccine awareness were recorded as < 3% and 0.5% respectively, in secondary school girls by Ezeanochie and Olasimbo [27]. While the findings in Figure 1 align with prior regional studies in SSA that describe similarly low baseline knowledge of HPV and the vaccine among both adults and adolescents [8,9], few studies have longitudinally tracked HPV awareness and action in mother-adolescent dyads during a national immunisation rollout.

Following the structured sensitisation introduced in April 2024 through the FACT study to stimulate awareness and action for both screening and vaccine uptake among mothers and adolescents, respectively, by leveraging the free, national HPV vaccine rollout. HPV awareness among mothers increased from 34.5 % at baseline to 64.4 % by the 12-month visit (p < 0.0001) as depicted in Figure 2, and vaccine awareness more than doubled (10.9 % to 27.6 %, p = 0.023). Cervical screening showed a similar pattern (Figure 3), while detailed proportions were provided in Table 3. VIA uptake rose from 35.6 % at the baseline to 49.7 % by the 12-month visit (p < 0.0001), whereas PAP smear rates remained static at ≈ 10 %. This aligns with WHO recommendations that prioritise VIA as a pragmatic first-line strategy in resource-limited settings and reflects the greater availability and immediate results advantage of VIA over cytology [28]. Nevertheless, overall screening coverage remains sub-optimal given Nigeria’s high cervical cancer burden [2,5].

Adolescent awareness also improved, albeit from a very low base, reaching 19% for HPV and 12.8% for the vaccine by the final visit. Vaccine uptake rose significantly from 0% at baseline to 3.4% by the 12-month visit (p < 0.0001), as observed in Table 4. The timing and magnitude of these gains (Figure 4) suggest that repeated community engagement and education were key drivers, consistent with evidence that structured outreach and school- or clinic-based education significantly improve HPV knowledge, screening, and vaccine acceptance in sub-Saharan Africa [1,18]. Study findings suggest that low-cost, embedded sensitisation in an HIV cohort can leverage national vaccine rollouts.

In Table 5, the most common reasons mothers cited for not screening were lack of symptoms (“haven’t had any problems,” > 60%) and absence of a provider recommendation (≈ 20 %). Our studies showed that healthcare provider endorsement was prominent both qualitatively and quantitatively and is among the strongest predictors of screening in low- and middle-income countries. These findings underscore missed opportunities in routine clinical encounters [14,17]. These results have immediate policy relevance as Nigeria scales up national HPV vaccination [11].

The absence of a significant association between HPV knowledge and sexual practices among mothers in Figure 5, even after further stratification by HIV status as shown in Figure 6, aligns with previous studies suggesting that knowledge acquisition alone does not necessarily translate into behavioural change [29]. While educational interventions improve understanding of HPV transmission and prevention, their impact on modifying intimate behaviours may be limited without accompanying strategies that address barriers to safer practices [30]. These findings therefore support the integration of behavioural and psychosocial components such as risk perception and supportive frameworks within HPV education programs targeted at mothers and other adult populations.

**Figure 5:**
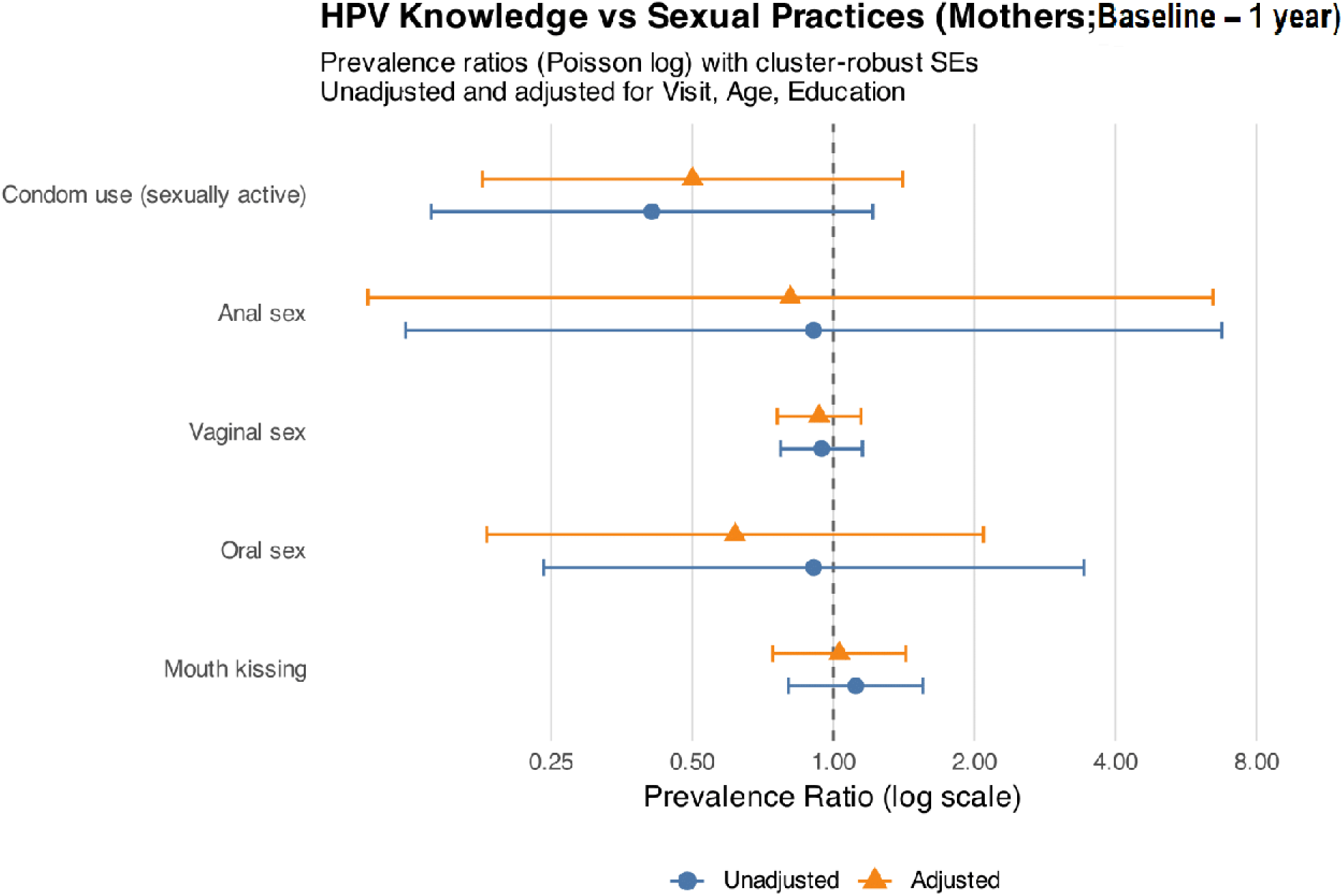
HPV knowledge did not impact sexual practices.

**Figure 6:**
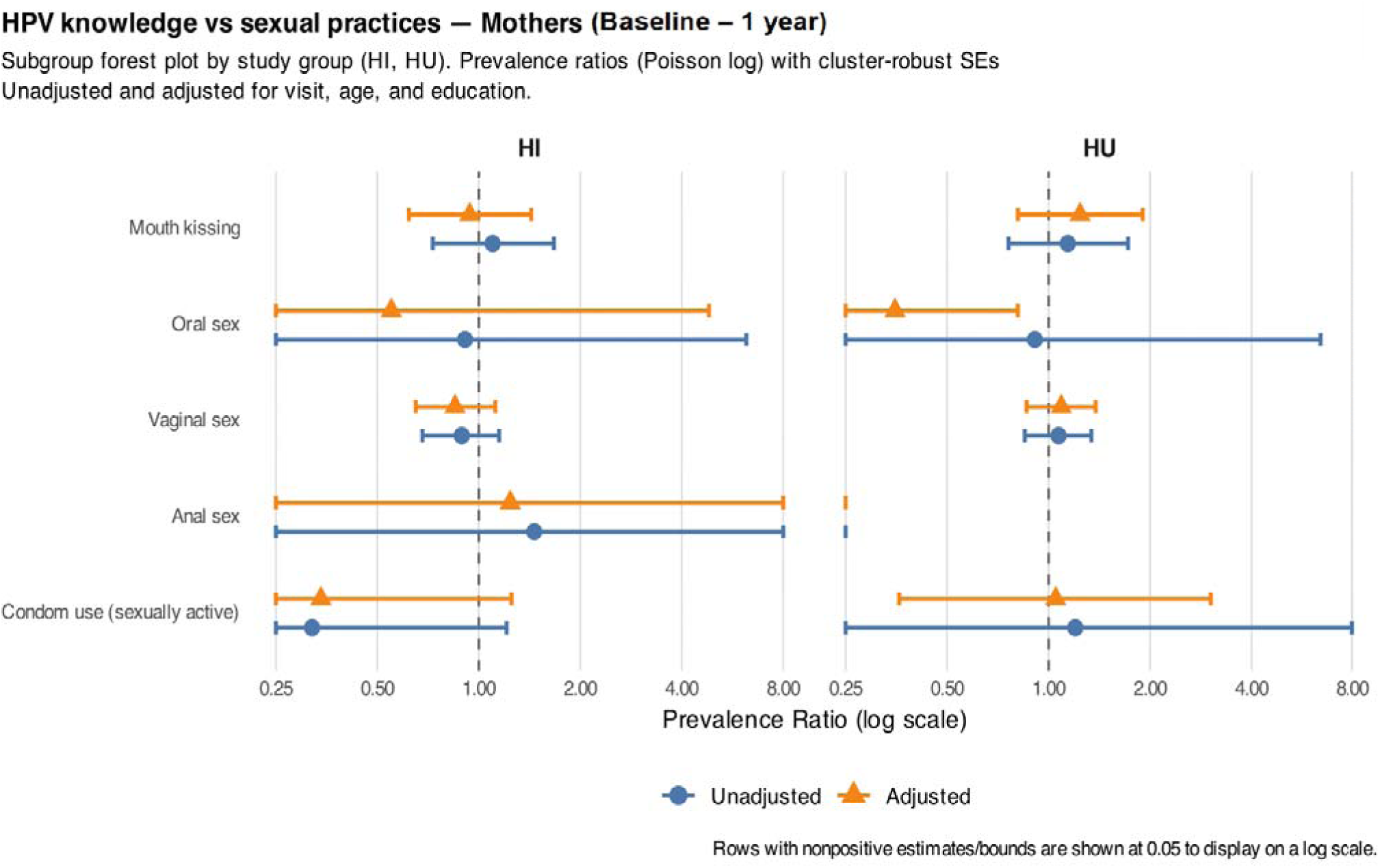
HPV knowledge did not impact sexual practices across HIV subgroups.

Qualitatively, participants’ responses show a gap between general awareness and specific knowledge, for example, that HPV causes cervical cancer, the vaccine is most effective at ages 9-14, or that screening can help early detection and prevent spread [1]. This pattern of reasonable surface awareness but poor depth of knowledge that limits preventive action has been reported elsewhere in Nigeria and the region. Studies have documented low detailed knowledge of HPV, cervical cancer, and prevention despite some awareness campaigns, and link poor knowledge to lower screening and vaccine uptake [31,32].

Many mothers expressed beliefs that they or their family are not susceptible to cancer or that screening is unnecessary in the absence of symptoms. Such misconceptions, including the idea that cervical cancer is not relevant unless symptoms are present, reduce preventive screening behaviour. Similar findings in Nigeria [33] and sub-Saharan Africa [30] have shown that low perceived susceptibility and erroneous beliefs undermine uptake of screening and vaccination. Fear about fertility effects, population control conspiracies, and memories of adverse events from other vaccine drives were prominent. These themes echo wider findings that vaccine mistrust, amplified by social media and pandemic-era conspiracy narratives, contribute to hesitancy for HPV vaccination in LMICs, including Nigeria, where parents may fear infertility or hidden agendas [30,35]. Systematic and qualitative reviews link conspiracy beliefs and mistrust to reduced vaccine confidence [14,37].

Quotes indicate that some women were told to “go and get tested” without a clear explanation of what the test was for or how HPV is transmitted. Poor provider communication and missed opportunities for education are common barriers identified; health care practitioners sometimes fail to explain the link between HPV and cancer or to counsel on prevention, leaving health seekers uncertain about the purpose and benefit of screening/vaccination [30,34]. Even when interest exists, participants asked practical questions about location, cost, turnaround time, and benefits. Thus, improving provider-patient communication and using clinic visits as teachable moments is recommended. Studies from Nigeria show that logistical barriers such as uncertainty about where services are offered, perceived or real costs, and convenience, meaningfully reduce uptake and must be addressed alongside information campaigns [33,34].

A notable strength of the FACT study was its integration into the HOMINY prospective cohort midway through implementation. Embedding FACT during three of the four scheduled clinical visits enabled the collection of detailed longitudinal quantitative data on sociodemographic factors, cervical screening, and vaccination status using standardised, interviewer-administered questionnaires to assess the impact. The qualitative component of the intervention sensitisation on HPV knowledge, screening, and vaccination provided a platform to evaluate participants’ reactions, learning, behavioural change, and outcomes.

Qualitative insights were strengthened by quantitative trends, allowing triangulation of findings and a nuanced understanding of the intervention’s impact over time. The convergence of these data builds confidence that the observed changes reflect genuine community-level behavioural shifts that translate awareness into action. Implementing FACT at this strategic midpoint enabled the team to measure baseline knowledge and behaviours, deliver targeted education just before Nigeria’s second-phase HPV vaccine rollout, and assess both immediate and longer-term effects on HPV knowledge and vaccine awareness in participant dyads, uptake among adolescents, as well as HPV screening among mothers.

However, findings rely on self-reported awareness and screening history, which may be affected by recall or social desirability bias. The study population may not be fully representative of all Nigerian communities, and generalisability is uncertain. Additionally, some subgroup analyses were underpowered. A much larger cohort across geopolitical zones in Nigeria will provide greater coverage and assessment.

To translate rising awareness into action, programmes should harness the channels that adolescents and their families already trust and use. Social media platforms and chatting apps popular with young people can host quizzes, short videos, and personal testimonials that dispel myths and reduce stigma around HPV vaccination and screening. Training peer educators in schools and adolescent clubs to lead debates, run interactive sessions, and share accurate information will further normalise preventive behaviours. At the same time, maternal influence can be reinforced by embedding HPV counseling and vaccination reminders into antenatal, postnatal, and routine child-immunisation visits, so mothers receive clear guidance on their own screening needs and their children’s eligibility for vaccination. Women’s associations, faith-based groups, and neighborhood meetings can amplify these messages, while outreach should explicitly include boys, highlighting that HPV is also a cause of oropharyngeal and anogenital cancer and that they, too, are susceptible and need vaccination.

Parallel efforts must focus on expanding accessible screening and treatment services. Training and equipping midwives, nurses, and community health workers to perform VIA and provide immediate treatment can rapidly extend coverage to underserved areas while gradually building laboratory capacity for HPV detection and cytology. This task-shifting approach brings preventive care closer to households, reducing travel barriers and increasing early detection rates. Integrating these clinical services with the community-driven education strategies creates a comprehensive, sustainable framework that protects women, men, boys, and girls from HPV-related cancers.

## Conclusion

HPV vaccine awareness and uptake in this Nigerian cohort were critically low at baseline among adolescents but improved substantially following targeted community-based sensitisation aligned with research activities and active follow-up of participants. VIA screening also increased significantly among mother-participants. These findings demonstrate that integrated, community-led interventions embedded within HIV care and research platforms can effectively promote STI prevention and contribute to broader cervical cancer elimination goals. Healthcare provider recommendations emerged as a key determinant of screening behaviour. To stimulate action for the national HPV vaccination rollout, a combined strategy will be essential, one that includes strong community education to raise awareness, address misconceptions and vaccine mistrust, active engagement of adolescents, and systematic prompts for healthcare providers. These components are critical to achieving the WHO’s targets for eliminating cervical and other HPV-related cancers. Additionally, improving access to HPV vaccination and scaling up screening among mothers will be vital for reducing the incidence of HPV-associated cancers.

## Data Availability

All data produced in the present study are available upon reasonable request to the authors

## Author Contributions

Conceptualisation; MOC, PA, EO^2^, YB, NS, NOP, OO, FEE-U

Data Curation; MOC, AK, RA, PA, EO^2^

Formal Analysis; EO, MOC, OP

Funding acquisition; MOC

Investigation; OP, EO^2^, PA, NLI

Methodology; OP, PA, EO^2^, NLI, MOC

Project Administration; MOC, OO, FEE-U, PA, EO^2^,

NLI Resources; PA, EO^2^, MOC

Software; MOC, PA, RA, AK

Supervision; MOC, PA, EO^2^,

Validation; MOC, PA, EO^2^, OP, AK

Visualisation; EO, MOC, OP

Writing (Original Draft Preparation); OP

Writing (Review and Editing); OP, MOC, PA, EO^2^, POO, NLI, EO, FEE-U.

## Acknowledgements

The authors are grateful to the women, children/adolescents, and families who made this study possible and to the dedicated study staff at the University of Benin Teaching Hospital and the Institute of Human Virology, Nigeria. Special thanks to all HOMINY Study Team members, including Lydia Esena-Obaweiki, Oluwaseun Peter, Promise Olumefun, Toluwalope Gbolahan, Isioma Mayor-Nwakwuribe, Alex Olanipekun, Amara Godwins, Osarobo Osadolor, Modupe Kadiri, Daniel Oakhu, Matthew Imoe-Auweokha, Jessica Kubeyinje, Daniel Anonyai, Erika Juhlin, Anil Kumar, and Dr. Uwagboe Odigie, for their invaluable contributions to participant engagement and recruitment, as well as sample and data collection and analysis. The authors also appreciate the dedicated efforts of the study dentists- Drs. Nneka Chukwumah, Stanley Iyorzor, Oseriemen Akhigbemen, Timothy Ahworegba, Gbenga Ojuola, and Sunday Adesiyan- for their time and expertise in conducting clinical examinations during study visits.

## Data Availability Statement

Data not publicly available; De-identified datasets generated in this study are not publicly available but can be provided upon request from the corresponding author.

## List of abbreviations

FACT: From awareness to action
HEU: adolescent HIV-exposed without acquisition
HI: = Individuals (adolescents or mothers) living with HIV
HOMINY: **H**uman Papillomavirus Human Immunodeficiency Virus, and **O**ral **M**icrobiota **I**nterplay in **N**igerian **Y**ouths
HPV: Human papillomavirus
HU: mothers HIV-unexposed
HUU: adolescent HIV-unexposed
LMICs: Low- and middle-income countries
OPSCC: Oropharyngeal squamous cell carcinoma
PAP: Papanicolaou
PLHIV: People living with
HIV SSA: sub-Saharan Africa
STIs: Sexually Transmitted Infections
VIA: Visual Inspection with Acetic Acid

